# Time to treatment initiation in pregnant women with tuberculosis in Cape Town, South Africa

**DOI:** 10.64898/2026.02.18.26346464

**Authors:** Sue-Ann Meehan, Anneke C. Hesseling, Emma Kalk, Jennifer A. Hughes, James A. Seddon, Victoria E. Namukuta, Muhammad Osman

## Abstract

**Background:** Tuberculosis (TB) incidence peaks in women during their reproductive years and is a leading cause of maternal mortality. Pregnant women with TB have a high risk of failure to initiate TB treatment and poor TB treatment and pregnancy outcomes. We determined the time to treatment initiation in pregnant women diagnosed with TB in a routine programmatic setting.

**Methods:** Using routine linked electronic data, we identified women 15-45 years of age with laboratory-confirmed and/or clinically diagnosed TB, October 2018-December 2020, in two high-burden sub-districts in Cape Town, South Africa. We compared demographic and clinical characteristics in women with TB by pregnancy status, used time-to-event analysis to determine the time from TB diagnosis to initiation of antituberculosis treatment and Cox regression to assess determinants of treatment initiation.

**Results:** Of 5,459 women diagnosed with TB, 292 (5.3%) were pregnant. The median age for pregnant women was 28.6 years (interquartile range [IQR]: 23.7–33.7) and non-pregnant women 31 years (IQR:25.2–36.5). HIV prevalence was similar in pregnant (177/292; 60.6%) vs non-pregnant (3200/5167; 61.9%) women. Median time to treatment initiation was two days for pregnant and non-pregnant women. Most women initiated treatment within the first month after their TB diagnosis, after which the rate plateaued in both groups. Time to treatment initiation over 6 months was statistically different (Kaplan Meier Log-rank test, p = 0.0064) with pregnant women lagging behind non-pregnant women.

**Conclusions:** More than 5% of women diagnosed with TB were pregnant at the time of TB diagnosis. While pregnant women with TB were appropriately initiated on treatment, almost 15% were never started on treatment and there were delays in treatment initiation. While strategic interventions to prioritise early treatment initiation are needed, there should be a specific focus on pregnant women who have not initiated treatment within one month after TB diagnosis.

## Background

Globally, an estimated 10.8 million people developed tuberculosis (TB) disease in 2023, of whom 3.6 million (33%) were women.^1^ TB incidence peaks in women during the reproductive years ^2-4^ and is a leading cause of mortality among women of childbearing age.^5^ Globally, an estimated 239,300 pregnant and 97,600 postpartum women developed TB in 2023.^6^ Pregnant women with TB have poor TB treatment outcomes^7 8^ and their babies are at high risk of premature birth, low birthweight and stillbirth.^8-11^ Failure to initiate TB treatment is a predictor of adverse pregnancy outcomes.^12^ Prompt initiation of TB treatment is therefore critical for all newly diagnosed individuals, especially pregnant women, to improve both TB treatment and maternal and neonatal outcomes.

South Africa is a high TB, TB-HIV and multidrug-resistant/rifampicin-resistant (MDR/RR)-TB burden country.^1^ In 2023, an estimated 270,000 people developed TB in South Africa, 37% were women ≥15 years, 5% had MDR/RR-TB and 54% of people with TB were living with HIV.^1^ Addressing TB requires an integrated approach that strengthens diagnosis, facilitates linkage to care with rapid initiation of appropriate treatment regimens, and supports treatment adherence. Early initiation of TB treatment is critical, as delays increase the risk of TB-related morbidity and mortality^10^ and contribute to ongoing transmission of *Mycobacterium tuberculosis (M*.*tb)*.^*13* 14^ An estimated 47,900 people with TB in 2023 in South Africa were not diagnosed.^1^ Between 12% and 30% of individuals diagnosed with TB in South Africa do not initiate treatment (“initial loss to follow-up”).^15 16^ Finding the missing people diagnosed with TB and linking them to care to start treatment is the central focus of the South African TB Recovery Plan initiated in 2022.^17 18^

In South Africa, time from TB diagnosis to TB treatment initiation varies between 0 and 144 days, depending on the diagnostic method used, and drug susceptibility test pattern of the infecting organism.^19^’^22^ Delays in treatment initiation among pregnant women are of particular concern, as they already face increased risks for both adverse TB and pregnancy outcomes. There is limited evidence on the time to TB treatment initiation in pregnant women with TB. To address this gap, we assessed the time from TB diagnosis to treatment initiation among pregnant women in a routine programmatic setting in South Africa.

## Methods

### Study design

This was a secondary analysis of programmatic data collected as part of a larger health system strengthening study (LINKEDin), which aimed to improve linkage to care following TB diagnosis.^15^

### Data collection

*We* utilized data extracted from the Western Cape Provincial Health Data Centre (PHDC), that included TB, HIV and pregnancy data. The PHDC is a comprehensive linked health information exchange platform in the Western Cape province and uses unique identifiers to integrate all electronic health data from routine health information systems in the public sector (laboratory, pharmacy, administrative and other clinical data) into single patient level records to improve patient care.^23^ The dataset included all people who were routinely diagnosed with TB (bacteriologically or clinically) at either a hospital or primary healthcare (PHC) facility in two high-burden health subdistricts (Khayelitsha and Tygerberg) in the Cape Metro, Western Cape Province, between October 2018 and December 2020. We included both drug-susceptible (DS) TB and drug-resistant (DR)TB, which included any resistance recorded. In South Africa, persons are notified with TB when they are registered in a TB treatment register i.e. when treatment is started at a PHC facility or a specialised TB treatment hospital.

For this analysis, we restricted the cohort to women between 15 and 45 years. We extracted demographic and clinical variables for TB by pregnancy status, date of TB diagnosis and date of TB treatment registration at a PHC facility or a specialized TB hospital. We defined time to treatment initiation as the number of days from date of TB diagnosis to the date of registration in a TB treatment register. For this analysis, we censored the data at 6 months after TB diagnosis.

### Statistical analysis

*We* compared characteristics of women with TB by pregnancy status. Descriptive statistics were used to describe baseline characteristics, the number and proportion of women starting TB treatment and median time to TB treatment initiation, by pregnancy status. We generated an inversion of the Kaplan-Meier curves to demonstrate the time from TB diagnosis to initiation of TB treatment, by pregnancy status. Cox regression was used to determine predictors of treatment initiation. Predictors were added incrementally, observing the change in significance of the likelihood ratio test of each model, to produce a final adjusted model. The relationship between predictors was considered and we avoided co-linearity in the final model. SAS software (version 9.4; SAS Institute, Inc, Cary, NC) was used for analysis.

### Ethical considerations

The Health Research Ethics Committee of Stellenbosch University (N18/07/069) approved the study, which was conducted according to the guiding principles within the Declaration of Helsinki. Approvals were received from the Western Cape Government Department of Health and Wellness and the City of Cape Town Health Directorate. We received a waiver of individual informed consent for this routinely collected deidentified health data.

## Results

### Overall

Between October 2018 and December 2020, a total of 5,459 women aged 15–45 years were diagnosed with TB; 292 (5%) were pregnant. Pregnant women had a median age of 29.0 years (interquartile range [IQR]: 24.0–34.2) and non-pregnant women were 31.3 years (IQR: 25.5–36.8). HIV co-infection was common, affecting 61.9% of all women, with similar prevalence across pregnancy status. The majority of women (70.8%) had bacteriologically confirmed disease. A higher proportion of pregnant women (39.7%) were diagnosed in hospital compared to those who were not pregnant (28.1%). The majority of the cohort (94%) had previously accessed care at a primary healthcare facility for any health services. (Table 1).

**Table 1:** Demographic and clinical characteristics of women with tuberculosis, by pregnancy status.

|  | Total (%) | Pregnant (%) | Not Pregnant (%) |
| --- | --- | --- | --- |
|  | (N=5459) | (n=292) | (n=5167) |
| **Age in years, median (IQR)** | 31.2 (IQR:25.3-36.6) | 29.0 (IQR:24.0-34.2) | 31.3 (25.5-36.8) |
| 15-19 | 518 (9.5) | 24 (8.2) | 494 (9.6) |
| 20-24 | 785 (14.4) | 61 (20.9) | 724 (14.0) |
| 25-29 | 1148 (21.0) | 78 (26.7) | 1070 (20.7) |
| 30-34 | 1293 (23.7) | 67 (22.9) | 1226 (23.7) |
| 35-39 | 980 (17.9) | 50 (17.1) | 930 (18.0) |
| 40-45 | 735 (13.5) | 12 (4.1) | 723 (14.0) |
| **HIV status** |  |  |  |
| Positive | 3377 (61.9) | 177 (60.6) | 3200 (61.9) |
| Negative | 2082 (38.1) | 115 (39.4) | 1967 (38.1) |
| **Place of TB diagnosis (%)** |  |  |  |
| Hospital | 1570 (28.8) | 116 (39.7) | 1454 (28.1) |
| PHC facility | 3889 (71.2) | 176 (60.3) | 3713 (71.9) |
| **TB diagnosis (%)** |  |  |  |
| Bacteriologically confirmed | 3865 (70.8) | 216 (73.9) | 3649 (70.6) |
| Clinically confirmed | 1594 (29.2) | 76 (26.1) | 1518 (29.4) |
| **Drug susceptibility** |  |  |  |
| DR-TB | 296 (5.4) | 14 (4.8) | 282 (5.5) |
| DS-TB | 3286 (60.2) | 190 (65.1) | 3096 (59.9) |
| DST not recorded | 1877 (34.4) | 88 (30.1) | 1789 (34.6) |
| **Site of disease** |  |  |  |
| Extra-pulmonary TB | 1018 (18.6) | 55 (18.8) | 963 (18.6) |
| *Pulmonary TB | 4108 (75.3) | 227 (77.7) | 3881 (75.1) |
| Not specified | 333 (6.1) | 10 (3.4) | 323 (6.3) |
| **\*\*Ever attended PHC before TB diagnosis** |  |  |  |
| Yes | 5137 (94.1) | 288 (98.6) | 4849 (93.8) |
| No | 322 (5.9) | 4 (1.4) | 318 (6.2) |
| **Treatment initiated** |  |  |  |
| Yes | 4676 (85.7) | 249 (85.3) | 4427 (85.7) |
| No | 783 (14.3) | 43 (14.7) | 740 (14.3) |

IQR: inter-quartile range, TB: tuberculosis; DR-TB: includes any resistance recorded; DS: drug-susceptible; DST: drug susceptibility test pattern; PHC: primary health clinic, \*Pulmonary TB (PTB) includes persons with PTB only and those with PTB and extrapulmonary TB (as per the WHO classification) \*\*Denotes if a patient had ever previously accessed a PHC facility for any health service

### Initiation ofTB treatment

The proportion of women who were initiated on treatment was similar between pregnant and non-pregnant women, with 249/292 (85.3%) of pregnant women and 4427/5167 (85.7%) of non-pregnant women initiating TB treatment. The median time to treatment initiation was similar; 2 days (IQR: 0–7 days) for pregnant women and 2 days (IQR: 0–6 days) for non-pregnant women. Most women initiated treatment within the first month after their TB diagnosis, after which the rate plateaued in both groups. The time to treatment initiation differed significantly by pregnancy status (Log-rank test, p = 0.0064) and was slower in pregnant women. See Figure 1 and Figure 2.

**Figure 1.**
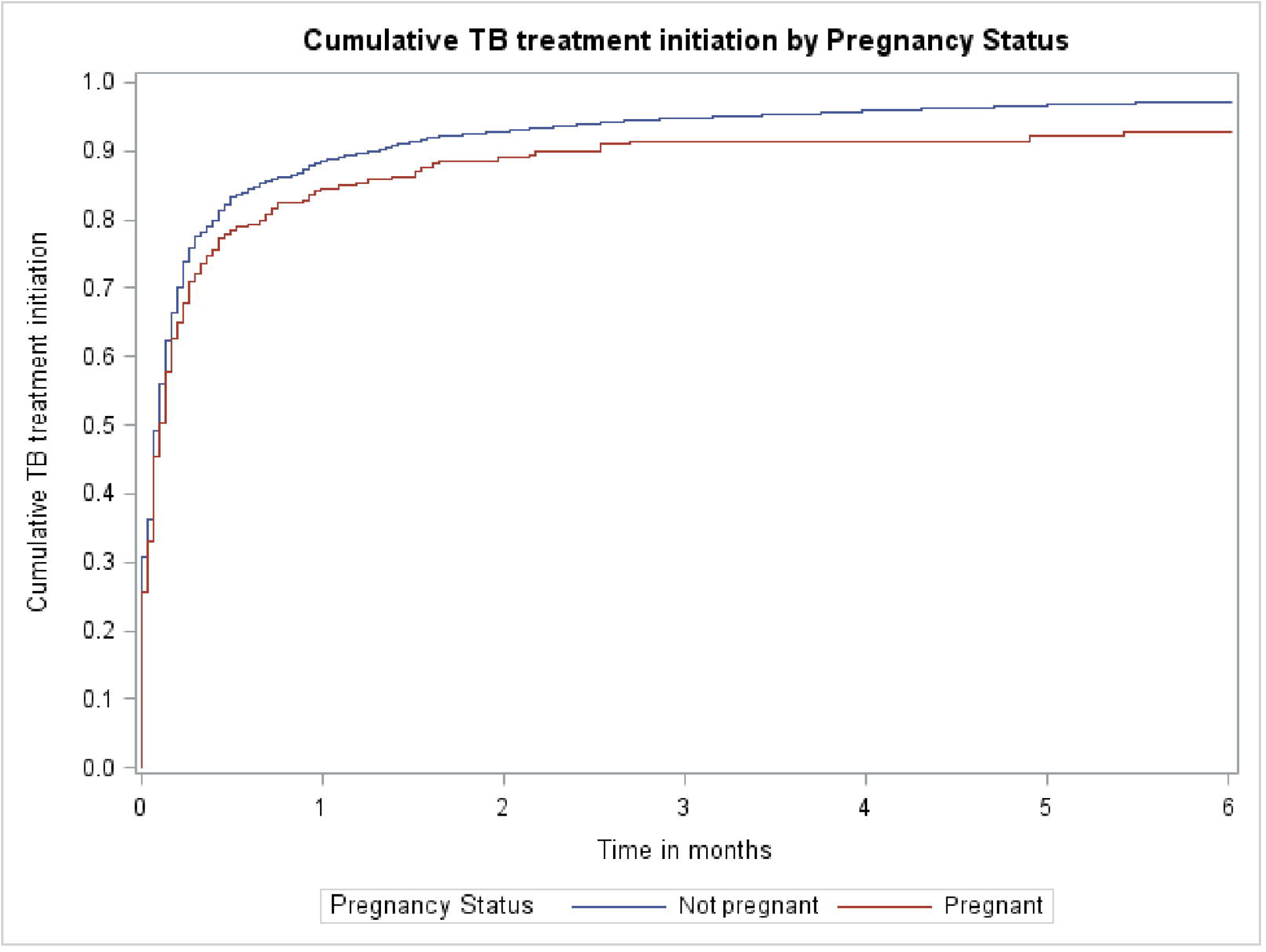
Time to treatment initiation stratified by pregnancy status over 6 months

**Figure 2.**
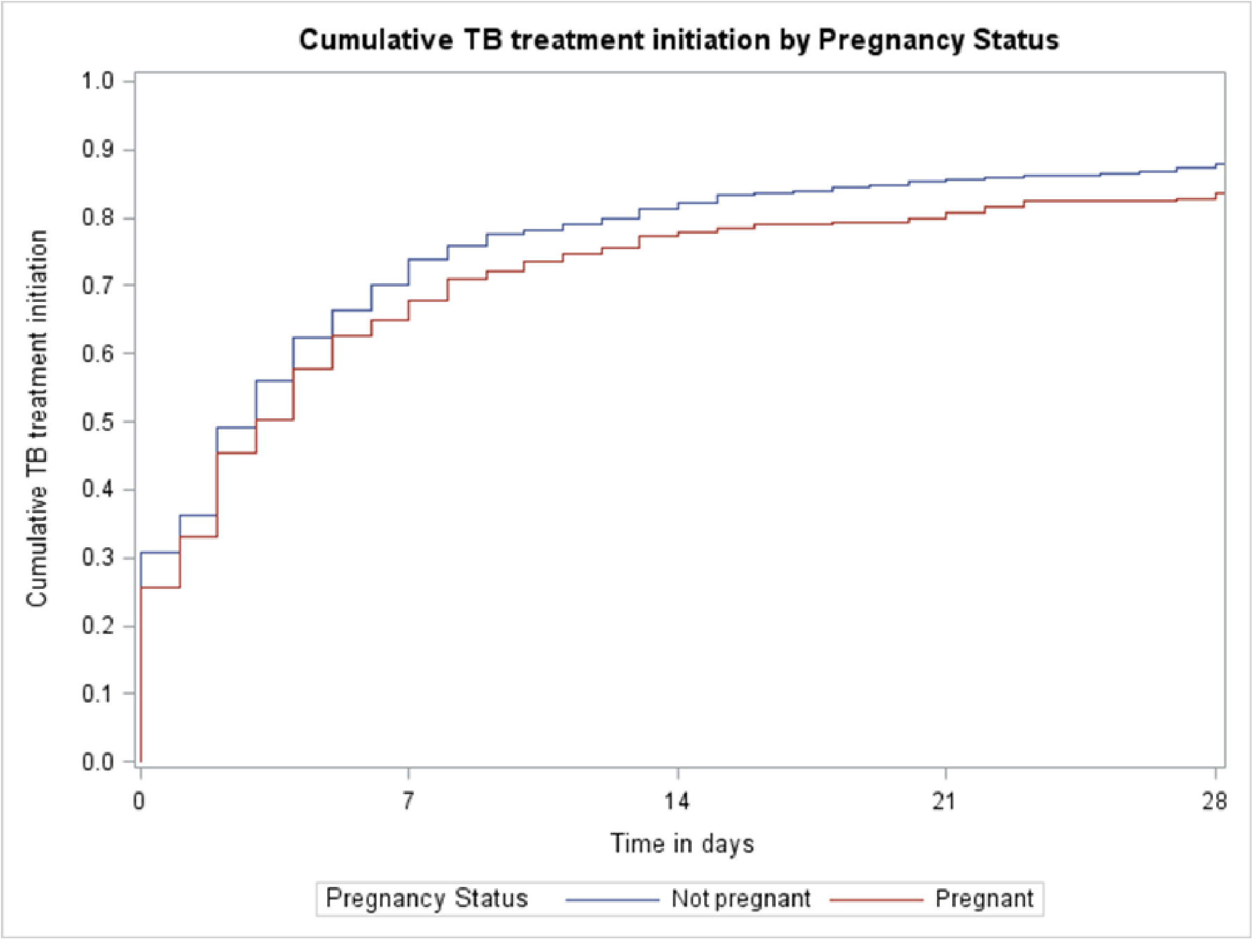
Time to treatment initiation stratified by pregnancy status in the first month following diagnosis

### Determinants ofTB treatment initiation

In multivariable analysis (Table 2), women who were HIV-negative had a slightly higher likelihood of treatment initiation compared to those living with HIV [adjusted hazard ratio (aHR) 1.08; 95% confidence interval (CI): 1.01–1.15]. Women who were diagnosed at a PHC facility compared to those diagnosed in hospital (aHR 3.86; 95% CI: 3.55–4.20) and women with a clinical TB diagnosis compared to having bacteriologically confirmed TB (aHR 4.98; 95% CI: 4.59–5.41) were significantly more likely to initiate treatment.

**Table 2:**
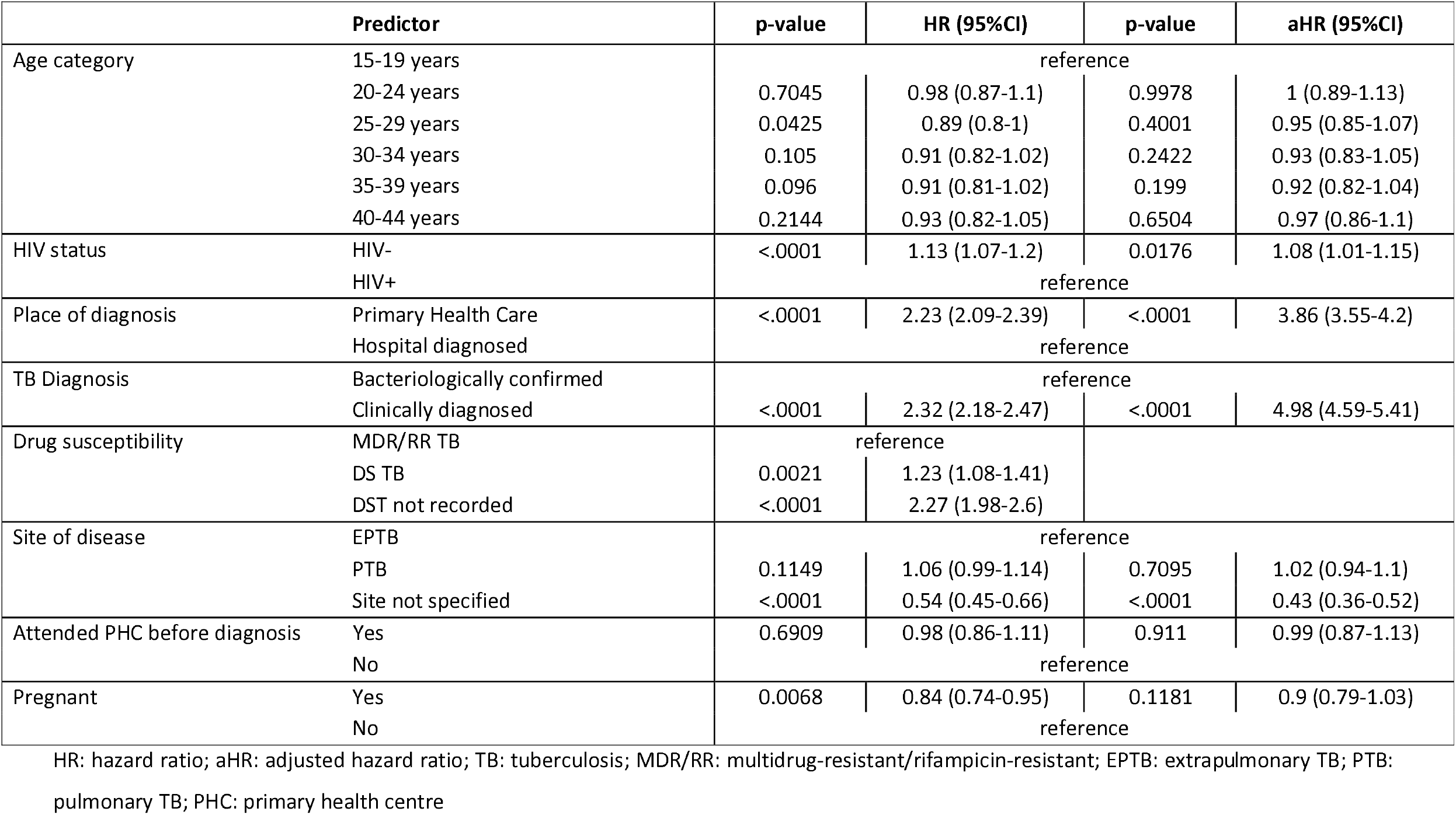
Predictors of treatment initiation among women 15-45 years (N=5459)

|  | Predictor | p-value | HR (95%CI) | p-value | aHR (95%CI) |
| --- | --- | --- | --- | --- | --- |
| Age category | 15-19 years | reference |  |  |  |
|  | 20-24 years | 0.7045 | 0.98 (0.87-1.1) | 0.9978 | 1 (0.89-1.13) |
|  | 25-29 years | 0.0425 | 0.89 (0.8-1) | 0.4001 | 0.95 (0.85-1.07) |
|  | 30-34 years | 0.105 | 0.91 (0.82-1.02) | 0.2422 | 0.93 (0.83-1.05) |
|  | 35-39 years | 0.096 | 0.91 (0.81-1.02) | 0.199 | 0.92 (0.82-1.04) |
|  | 40-44 years | 0.2144 | 0.93 (0.82-1.05) | 0.6504 | 0.97 (0.86-1.1) |
| HIV status | HIV- | <.0001 | 1.13 (1.07-1.2) | 0.0176 | 1.08 (1.01-1.15) |
|  | HIV+ | reference |  |  |  |
| Place of diagnosis | Primary Health Care | <.0001 | 2.23 (2.09-2.39) | <.0001 | 3.86 (3.55-4.2) |
|  | Hospital diagnosed | reference |  |  |  |
| TB Diagnosis | Bacteriologically confirmed | reference |  |  |  |
|  | Clinically diagnosed | <.0001 | 2.32 (2.18-2.47) | <.0001 | 4.98 (4.59-5.41) |
| Drug susceptibility | MDR/RR TB | reference |  |  |  |
|  | DS TB | 0.0021 | 1.23 (1.08-1.41) |  |  |
|  | DST not recorded | <.0001 | 2.27 (1.98-2.6) |  |  |
| Site of disease | EPTB | reference |  |  |  |
|  | PTB | 0.1149 | 1.06 (0.99-1.14) | 0.7095 | 1.02 (0.94-1.1) |
|  | Site not specified | <.0001 | 0.54 (0.45-0.66) | <.0001 | 0.43 (0.36-0.52) |
| Attended PHC before diagnosis | Yes | 0.6909 | 0.98 (0.86-1.11) | 0.911 | 0.99 (0.87-1.13) |
|  | No | reference |  |  |  |
| Pregnant | Yes | 0.0068 | 0.84 (0.74-0.95) | 0.1181 | 0.9 (0.79-1.03) |
|  | No | reference |  |  |  |

HR: hazard ratio; aHR: adjusted hazard ratio; TB: tuberculosis; MDR/RR: multidrug-resistant/rifampicin-resistant; EPTB: extrapulmonary TB; PTB:
pulmonary TB; PHC: primary health centre

In univariable analysis, pregnant women were significantly less likely to initiate treatment compared to non-pregnant women (HR 0.84; 95% CI: 0.74–0.95). However, after adjusting for age, HIV status, site of diagnosis, and method of TB diagnosis, pregnancy was no longer significantly associated with treatment initiation status (aHR 0.90; 95% CI: 0.79–1.03) (Table 2).

## Discussion

This study determined time to treatment initiation for pregnant and non-pregnant women diagnosed with TB within a routine programmatic setting in two sub-districts in South Africa, thereby addressing the evidence gap on time to treatment initiation among pregnant women with TB. We found that 85% of women started treatment, irrespective of pregnancy status, however pregnant women demonstrated slightly longer time to treatment initiation and ultimately the proportion starting treatment did not reach the same level as non-pregnant women within six months. This is an important finding, indicating that pregnant women with TB need additional support linking to care following diagnosis.

In South Africa, time to treatment initiation varies by setting, population group, TB type, and presence of comorbidities. In our cohort, the proportion of women starting treatment following TB diagnosis, was comparable to that reported among pregnant women in the same setting^12^ and similar to hospital-diagnosed patients in rural South Africa.^24^ However, it was higher than the treatment initiation rates observed among TB patients diagnosed in an urban South African hospital,^25^ MDR/RR-DR-TB patients,^21^ and individuals living with HIV who were not on antiretroviral therapy (ART).^26^

While median time to treatment initiation was the same in pregnant and non-pregnant women (2 days), time to treatment initiation over 6 months was statistically different with pregnant women lagging behind non-pregnant women. The South African National Department of Health (DoH) recommends that people diagnosed with DS-TB should initiate treatment within 2 days and those diagnosed with DR-TB, within 5 days.^27^ In our urban setting in Cape Town, pregnant women were able to access treatment within the targets set by the DoH for all patients diagnosed with TB. However, it is concerning that 30% of pregnant women and 25% of non-pregnant women had not started TB treatment within 7 days. Any health system interventions to improve treatment initiation rates in general (and time to treatment after 7 days) should include and prioritise pregnant women who appear to take much longer to initiate treatment than non-pregnant women. A systematic review reported a median delay of 14 days (95% CI: 3-84) between diagnosis and treatment initiation in low- and middle-income countries.^28^ Two South African studies reported the median time to treatment was 0 days in an urban setting, when Xpert MTB/RIF was used as the diagnostic test.^19 22^ Another study reported 14 days in a rural setting.^20^ While median time to treatment in our study was short for both pregnant and non-pregnant women, explaining the lag experienced by pregnant women is more complex and may be driven by patient delays in health seeking and/or health system challenges.

The proportion of women living with HIV in this cohort was high (62%), regardless of pregnancy status and exceeded the 2023 estimate where half of all individuals with TB (54%) in South Africa were co-infected with HIV^1^. The high HIV prevalence within our cohort indicates a substantial burden of disease, where most women are simultaneously navigating TB, HIV, and reproductive health services. In South Africa, public antenatal services are not routinely integrated with other primary healthcare services, while TB and HIV services are integrated to varying extents.^29 30^ It is probable that women may have attended antenatal services in the period between diagnosis and treatment initiation. This fragmentation in care delivery may contribute to delays in initiating TB treatment among women who must access multiple, disconnected healthcare services. In addition, pill burden could also explain delays in initiating TB treatment by overwhelming women who are living with HIV and now have the added burden of additional pills and regimens.^31 32^

Overall, nearly a third of women in our cohort were diagnosed with TB in hospital, with a notably higher proportion among pregnant women compared to non-pregnant women. The majority of this cohort had TB that was bacteriologically confirmed but this was similar by pregnancy status. A hospital-based diagnosis may indicate delayed recognition of TB symptoms. Having a negative Xpert MTB/RIF *(M*.*tb* nucleic acid amplification test) test result on initial testing can reflect more advanced disease. Pregnancy can complicate TB diagnosis, as physiological changes and overlapping symptoms may mask classic TB presentations. Healthcare providers are also hesistant to perform chest X-rays on pregnant women.^34^ This diagnostic complexity can contribute to delays, which in turn increase the risk of adverse maternal, pregnancy, and neonatal outcomes. It is acknowledged that time to treatment initiation for women diagnosed with TB in hospital may be overestimated, as some women may have initiated TB treatment during their hospital stay. However, any treatment initiated in hospital or at discharge that was not captured in the electronic data feeding into the PHDC would not have been included in our dataset. It is important to note that registration for treatment initiation at the primary healthcare (PHC) level typically includes essential counselling and adherence support, which are critical for successful treatment outcomes. Although we were unable to determine whether some individuals may have initiated TB treatment at a hospital, registration and initiation at a PHC facility represent the optimal pathway for ensuring continuity of care and adherence support.

Given that national guidelines recommend evaluation for TB in pregnant women at all antenatal visits,^35^ and targeted universal TB testing in pregnancy^36^, it is expected that pregnant women would be more likely to be tested for TB. At the same time, pregnant women may face competing health priorities and, where TB screening and testing requires an additional contact point, it may not be prioritised by them. There may also be a lack of awareness among women regarding TB symptoms and when to seek help. Lack of knowledge has been shown to be a predictor of treatment delays.^37 38^ This highlights the need for strengthened community-level education and awareness initiatives as a critical intervention to improve early recognition of TB symptoms and promote timely health-seeking behavior, particularly among pregnant women. Increasing the demand for TB screening and testing by women of reproductive age and especially pregnant women is an important consideration for public health services.

Being pregnant was associated with a lower likelihood of initiating treatment but was not significant in the multivariable analysis. This suggests that the initially observed association may be confounded by other clinical (e.g. disease severity or comorbidities), or healthcare system (e.g. access to care, provider decision-making) factors. As the effect of pregnancy reduced in the multivariable model, the adjusted hazard ratio for site or mechanism of diagnosis almost doubled, highlighting the need to prioritise diagnosis at PHC and where possible using TB-NAAT testing. This finding highlights the need to examine the broader healthcare context to reduce TB treatment delays as structural or clinical complexities may influence care pathways and linkage beyond individual characteristics such as pregnancy.

The major strength of this study is that it fills a critical gap in the scientific literature. While treatment delays among TB patients are well documented, there are limited data on time to treatment initiation among pregnant women with TB. As the study used routine data, it demonstrates a mechanism for national TB programmes to evaluate time between diagnosis and treatment registration which may be used as an estimate for actual treatment initiation. In addition, this study provides crucial linkage to TB care data for women in their reproductive years, an under-researched group identified by the WHO, as facing unique health risks and challenges. These risks have significant implications for maternal, child, and public health. Failing to disaggregate data by sex and age represents a missed opportunity to understand and address the specific needs of this population, which is essential for developing better, safer, and more equitable TB care.

Using linked electronic health data curated by the Provincial Health Data Centre (PHDC), enabled the use of routine programmatic data from multiple integrated sources, that allowed us to evaluate time to treatment initiation for pregnant and non-pregnant women with TB in a programmatic setting.

The study had several limitations. First, the study made use of routine electronic data which was limited by the variables available. While we attempted to consider those factors affecting treatment initiation, no information on the broader social determinants of health were available. Time to treatment initiation will need further exploration including understanding the individual context and perspective. Second, missing or incorrect data leading to misclassification must be considered. Where women attended TB services or where women received TB treatment in hospital but recording in the electronic systems was incomplete, we may have over estimated the treatment delays. Third, in our dataset, the first available evidence of a clinical TB diagnosis is often the recorded date of treatment initiation. Consequently, for women who were clinically diagnosed with TB, the date of diagnosis and treatment initiation are frequently the same, which may limit the ability to distinguish between the timing of diagnosis and initiation of treatment. Fourth, we did not investigate TB and treatment initiation of women during the postpartum period, where the risk of TB is substantial.

While this study demonstrates good overall linkage to TB care and TB treatment registration, the time taken to start treatment in some women is concerning. Since we used linked routine data, time-related elements are already available in the PHDC, and consideration should be given to introducing alerts and reporting flags for all individuals diagnosed with TB. Prioritizing pregnant women could help ensure that opportunities for linkage to care after screening and diagnosis are not missed during future health service visits. Further work is needed to investigate how time-to-treatment initiation indicators and integration with existing community outreach services can improve timely access to TB care.

## Conclusion

While pregnant South African women with TB largely started treatment, almost 15% were never started on treatment and delays in initiating TB treatment persist. Ongoing transmission risks, and increased morbidity and mortality for mother and infant require more strategic interventions to prioritise early treatment initiation. There should be a specific focus on pregnant women who have not initiated treatment within one month after TB diagnosis. Women diagnosed in hospital or bacteriologically are at especially high risk, and use of available digital tools should be considered to limit missed opportunities for treatment initiation during other contacts with health services

## Data Availability

The data are not freely available as they are secured by the WCGHW and application can be made to the WCGHW for access

## Acknowledgements

We wish to acknowledge staff at the Centre for Integrated Data and Epidemiological Research (CIDER) at the University of Cape Town and staff at the Western Cape Provincial Health Data Centre (PHDC) for their invaluable assistance, especially Mariette Smith and Florence Phelanyane, who were instrumental in assisting with the TB, HIV and pregnancy cascades. We are thankful to Dr Rory Dunbar at Stellenbosch University, for his data management expertise.

## Contributorship statement

SM and MO planned and designed the study. MO did the data extraction and produced the analysis. SM and MO interpreted the data. SM wrote the first draft and finalised the submission based on author and peer reviewer input.

All authors provided critical input into the manuscript, have reviewed the final version of the manuscript and approve of its content and submission for publication.

## Ethical statement

Stellenbosch University Health Reseach Ethics Committee (N18/07/069). The study included routine programmatic data and so did not include human participants directly.

